# Bottom-up and top-down contributions to impaired motion processing in schizophrenia

**DOI:** 10.1101/2023.07.07.23292259

**Authors:** Antígona Martínez, Pablo A. Gaspar, Dalton H. Bermudez, M. Belen Aburto-Ponce, Daniel C. Javitt

**Author notes:** Corresponding Author: Antígona Martínez Nathan Kline Institute for Psychiatric Research 140 Old Orangeburg Road, Orangeburg, NY 10962, USA (voice).

## Abstract

**Background and Hypothesis:** Motion processing deficits in schizophrenia have been linked to impairments in higher-order social-cognitive processes. The neural underpinnings are not fully understood but it has been hypothesized that middle temporal area (MT+) may serve as a bridge between purely sensory and more cognitive proceseses. We investigated the interrelationship between MT+ sensory processing deficits and impairments in higher-order processing using naturalistic videos with explicit motion and static images with implied-motion cues.

**Study Design:** Functional magnetic resonance imaging was used to evaluate cortical and subcortical brain regions associated with real- and implied-motion processing in 28 individuals with schizophrenia and 20 neurotypical controls. These measures were related to face emotion recognition and motion-perception deficits, as measured behaviorally.

**Study Results:** Activation of MT+ was abnormal in schizophrenia during both real- and implied-motion processing. Dysfunction of early visual cortex and pulvinar were also associated with impaired real-motion processing. During implied-motion-perception, MT+ participated in a wider network involving sensorimotor and prefrontal nodes of the human mirror neuron system, known to play a role in social-cognitive processes. Perception of both real- and implied-motion engaged the posterior superior temporal sulcus, a key node of the social brain network.

**Conclusions:** The findings support concepts of MT+ as a bridge between visual sensory areas and higher-order brain regions especially in relationship to face emotion recognition and social cognition. Our data argue for greater focus on MT+ contributions to social-cognitive processing, in addition to its well-documented role in visual motion processing.

## Introduction

Schizophrenia (Sz) is a serious mental disorder associated with a range of cognitive impairments predictive of long-term functional and social outcome (Green et al. 2019). The underlying causes are multifactorial, nevertheless, over recent years dysfunctional auditory and visual sensory processing have become increasingly appreciated as a hallmark of Sz and important contributors to higher-order cognitive impairments (Javitt and Freedman 2015). In the visual domain impaired motion-perception (e.g. reduced detection sensitivity of moving stimuli) has been linked to impairments in social-cognitive processes including theory of mind (TOM) (Kelemen et al. 2005), biological-motion perception (Okruszek and Pilecka 2017) and face emotion recognition (FER) (Martinez et al. 2018; Martinez et al. 2019). These findings are consistent with the recent proposal of the existence of a specific visual pathway specialized for social perception (Pitcher and Ungerleider 2021) in which motion is a fundamental component. This ‘third visual pathway’, located at the lateral surface of the brain, runs from primary visual cortex (V1) through visual motion-processing regions located in middle temporal cortex, to the superior temporal sulcus (STS), an area known to respond to a variety of dynamic social cues including faces and bodies (Allison et al. 2000; LaBar et al. 2003).

In neurotypical (NT) individuals, motion-perception engages an interconnected network of subcortical and cortical areas that converge on the middle temporal complex (MT+), which comprises the motion processing center of the primate brain (Maunsell et al. 1990). Subcortical (thalamic) structures include the magnocellular-recipient layers of the lateral geniculate nucleus (LGN), and the lateral and inferior divisions of the pulvinar nucleus (PulN). MT+ also receives input from deep layers of V1 (Movshon and Newsome 1996). Functional magnetic resonance imaging (fMRI) studies have shown that MT+ activation is reduced in Sz in response to simple moving stimuli, in parallel with elevated motion-detection thresholds and reduced motion-induced event-related potentials (ERP) (Chen et al. 2008; Lencer et al. 2005; Martinez et al. 2018; Martinez et al. 2019). Additionally, MT+ activation deficits in Sz are associated with aberrant interactions involving V1 and PulN (Martinez et al. 2018; Martínez et al. 2022).

The sensitivity of MT+ is not limited to real physical motion but extends also to the representation of *implied-motion* derived from static images containing cues conveying information about an objects’ motion direction and speed (Fawcett et al. 2007; Kourtzi and Kanwisher 2000; Osaka et al. 2010; Senior et al. 2000; Zeki et al. 1993). The involvement of MT+ during implied-motion perception has been attributed to activity of the mirror neuron system (MNS) (Filimon et al. 2007; Proverbio et al. 2009; Urgesi et al. 2006). Mirror neurons, originally discovered in macaques, discharge during both active execution and passive observation of a motor act, thereby “mirroring” the goal-directed actions of others (reviewed in Bonini et al. 2022).

FMRI studies have consistently identified a set of ‘core’ regions comprising the human MNS such as the premotor, inferior prefrontal and parietal cortices (Caspers et al. 2010; Molenberghs et al. 2012), along with a more extended network of cortical areas that include the sensorimotor cortex, supplementary motor area, posterior superior temporal sulcus (pSTS), MT+, and putamen, (Bonini 2017; Caspers et al. 2010; Errante and Fogassi 2020; Ferrari et al. 2017). The MNS also plays a central role in social cognitive processes including TOM, empathy, and FER (Schmidt et al. 2021). Dysfunctional MNS activity has been associated with social cognitive impairments across neuropsychiatric disorders (reviewed in Chan and Han 2020; Jeon and Lee 2018). In Sz, studies have reported reduced MNS activation during action observation (Mehta et al. 2014; Valizadeh et al. 2022), in association with impaired TOM performance (Choe et al. 2018) and negative symptom severity (Schilbach et al. 2016).

In recent studies (Martinez et al. 2018; Martinez et al. 2019; Martínez et al. 2022), we have observed reduced MT+ activation in Sz in response to simple motion stimuli which correlate with reduced FER to static emotional faces (Martinez et al. 2018), suggesting a potential involvement of MT+ deficits even to static-only stimuli.

In this study, we investigated the parallel cortical circuitry underlying the perception of real- and implied-motion in individuals with Sz. We used naturalistic stimuli that either contained real-motion (videos) or in which motion, or its absence, were implied by static photographs. Our goal was to investigate whether MT+ deficits to explicit motion in Sz would carry forward even to the processing of static, implied-motion stimuli and, if so, whether the MT+ deficits would mediate impairments in activation of higher-tier social cognitive regions, such as pSTS.

We hypothesized that MT+ and pSTS would show impaired activation to both real- and implied-motion stimuli, and that significant correlations would be observed across stimulus types, arguing for intrinsic dysfunction within the third visual pathway system. Moreover, based on our previous studies (Martinez et al. 2019; Martínez et al. 2022), we hypothesized that MT+ deficits in Sz to explicit (real) motion would be associated with impaired sensory (“bottom-up”) processing whereas deficits in response to stimuli with implied-motion would be associated with (“top-down”) dysfunction of high-order cortical areas.

Finally, while the majority of studies of cognitive impairment in Sz focus on cortical activations, it is increasingly appreciated that the network structure of the brain is coordinated extensively through subcortical regions including thalamus and basal ganglia, which participate in canonical resting-state networks (Choi et al.; Ji et al. 2019; Zhang et al. 2008). Thus, in the present study we additionally investigated functioning of subcortical regions implicated in real- and implied-motion processing.

## Materials and Methods

### Participants

Participants were 28 individuals meeting DSM-IV criteria for Sz assessed by the Structured Clinical Interview for DSM-IV (SCID) (First et al. 1997) and 20 demographically-matched NT controls with no history of SCID-defined axis I psychiatric disorders. Controls were recruited from the volunteer recruitment pool at the Nathan Kline Institute for Psychiatric Research (NKI). Sz participants were recruited from outpatient and chronic inpatient clinics in the New York City area and were on stable doses of antipsychotics at time of testing. Chlorpromazine-equivalents (CPZ-equivalents) were estimated as described previously (Andreasen et al. 2010) **(Table 1**). Participants were excluded if they had neurological or ophthalmologic disorders or if they met criteria for alcohol/substance dependence within the last 6 months or alcohol/substance abuse within the last month. The investigation was approved by the NKI institutional review board. Informed consent was obtained from all participants after study procedures were fully explained. In Sz, symptoms were assessed using the Positive and Negative Symptom Scale (PANSS). Neurocognitive performance was assessed using the Processing Speed Index (PSI) and Perceptual Organization Index (POI) of the WAIS-4.

**Table 1:** Demographic, clinical and behavioral characteristics. Neurotypical (NT), schizophrenia (SZ) participants differed in terms of years of education, socioeconomic status (SES), Processing Speed Index (PSI) and Perceptual Organization Index (POI). Additionally, face-emotion recognition scores, derived from the Penn Emotion Recognition Task (ER-40) were significantly lower in the SZ group as well as motion sensitivity, calculated as 1/threshold on the coherent motion detection task. CPZ: Chlorpromazine equivalents; PANSS: Positive and Negative Syndrome Scale. Standard deviations in parentheses.

|  | **NT (n=20)** | **SZ (n=28)** | **Difference** |
| --- | --- | --- | --- |
| **Age** | 35.5 (9.4) | 38.8 (10.1) | t(46)=-1.15, p=.25 |
| **Sex (F/M)** | 4/16 | 8/20 | $\chi^2=.45$, p=.49 |
| **IQ** | 99.9 (8.3) | 98.1 (9.3) | t(46)=.69, p=.49 |
| **Years of Education** | 14.7 (2.1) | 11.9 (1.9) | t(46)=4.76, p<.001 |
| **Participant SES** | 42.7 (10.9) | 25.0 (6.2) | t(46)=7.11, p<.001 |
| **Parental SES** | 42.5 (14.4) | 37.5 (12.7) | t(46)=1.17, p=.25 |
| **PSI** | 100.1 (11.0) | 83.7 (11.3) | t(46)=4.89, p<.001 |
| **POI** | 108.9 (15.9) | 93.4 (18.3) | t(46)=2.98, p=.005 |
| **Illness Duration (yrs)** | -- | 14.2 (9.1) | -- |
| **Edinburgh Score** | 15.7 (6.6) | 16.9 (5.4) | t(46)=-.69, p=.49 |
| **CPZ Equiv. SZ)** | -- | 834.8 (629.1) | -- |
| **Anti-Psychotic** |  | Typical (6) | -- |
|  |  | Atypical (15) | -- |
|  |  | Combination (7) | -- |
| **ER-40** | 35.8 (1.9) | 28.7 (5.6) | t(46)=5.56, p<.001 |
| **Motion Sensitivity** | 10.9 (4.2) | 7.3 (3.0) | t(46)=3.41, p=.001 |
| **PANSS (positive)** | -- | 11.1 (4.1) | -- |
| **PANSS (negative)** | -- | 19.3 (5.6) | -- |

The Penn Emotion Recognition (ER-40) test (Taylor et al. 2012) was administered to all participants to ascertain FER accuracy.

Individual thresholds for coherent-motion detection were determined using random-dot kinematograms as described previously (Martinez et al. 2018). Analyses were carried out on motion-sensitivity scores, calculated as 1 over the estimated coherence threshold for each participant.

### Task Design

Stimuli depicted animals in the wild or humans performing a sports activity. For each category, three stimulus types were used: 1) black and white video clips containing real-motion; 2) greyscale photographs depicting implied-motion; and 3) greyscale photographs depicting motion-absence. Implied-motion stimuli were single frames captured from real-motion videos. No-motion images were chosen to match real- and implied-motion stimuli. All images were matched for luminance and contrast using the SHINE toolbox (Willenbockel et al. 2010).

Stimuli were delivered in 24-second blocks. During real-motion blocks, eight unique 3-second video clips were presented with no break between clips. In implied- and no-motion blocks, eight unique images were presented for 3-seconds each. Stimulation blocks were interleaved with 12-seconds of fixation alone. A total of 12 blocks (real-, implied- or no-motion) from each category (sports or animals) were delivered in random order in each of two separate fMRI scans. Participants monitored a central fixation cross and responded by button-press upon detection of its occasional dimming occurring every 3-9 seconds.

### Data acquisition and pre-processing

FMRI data were collected on a Siemens 3T TIM-Trio scanner. At least one high-resolution structural image was acquired per participant using a Magnetization-Prepared Rapid Gradient-Echo (MP-RAGE) sequence (spatial resolution=1mm isotropic, repetition time (TR)=2000ms; echo time (TE)=3.5ms; flip angle 8°, 192 slices). Echo-planar images (EPIs) (2.5mm isotropic, TR=2000ms, TE=30ms, flip angle 80°) were acquired on 36 contiguous slices in the axial plane.

Functional data were preprocessed and analyzed using a combination of Analysis of Functional NeuroImages (AFNI) (Cox 1996), Surface Mapping (SUMA) (Saad and Reynolds 2012). Preprocessing consisted of concatenating data from two runs, removal of signal deviation >2.5 SDs from the mean (3dDespike), temporal alignment, identification of motion outliers per run and scaling of blood-oxygen-level-dependent (BOLD) values to mean percent signal change (Taylor et al. 2018). Cortical data was spatially smoothed with a 6mm full-width-at-half-maximum Gaussian kernel.

For each participant, the cortical surface was rendered using FreeSurfer’s (Version 7.2.0, FreeSurfer.net) ‘reconall’ command and standard preprocessing steps including motion correction, removal of non-brain tissue, skull-stripping, segmentation of white and gray matter and intensity normalization.

Analyses were carried out on the gray-matter ordinates of each individual cortical surface aligned to the FreeSurfer (fsaverage)141-standard mesh. Cortical data was sampled to the Human Connectome Project multimodal cortical parcellation atlas (HCP-MMP1.0) (Glasser et al. 2016), resampled to fsaverage. The HCP atlas subdivides the brain into 180 regions (parcels) per hemisphere based on functional and structural properties. For subcortical structures, parallel analyses were carried out in volumetric space registered to the MNI-152 template brain.

### fMRI data analysis

A general linear model (GLM) approach was used to estimate beta parameter values across the whole brain for each stimulus type (real-, implied-, no-motion). Regressors were constructed by convolving a gamma-variate function with a boxcar function representing the timing of each stimulation block type. Additionally, six motion parameters (three translation and three rotation parameters per frame) as well as white matter (WM) and cerebrospinal fluid (CSF) signal time series were included in GLM as nuisance effects.

Maps of beta parameter estimates for the contrast of activation during real-versus no-motion and implied-versus no-motion stimulus blocks were created for individual participants in separate general linear tests (GLT).

To avoid issues related to circularity in data analysis (Kriegeskorte et al. 2009), whole-brain contrast maps for each condition were first analyzed by analysis of variance (ANOVA) across all participants. Monte Carlo simulations (*slow_surf_clustsim.py)* were run to estimate the surface area cluster size associated with a family-wise error rate of α<.05. The resultant groupwise maps were used to identify parcels for use in subsequent between-group analyses.

From each parcel identified in the groupwise analyses, mean beta parameter estimates for the real- and implied-motion contrasts were extracted and entered into repeated-measures ANOVA with a between-group factor of group membership (NT, Sz) and within-group factor of parcel.

In addition, beta values were extracted from two a-priori identified subcortical regions of interest (ROIs), the pulvinar nucleus of the thalamus and basal ganglia (caudate, putamen and globus pallidus) identified anatomically using a template mask in standard MNI space.

Univariate tests were used for post-hoc comparisons of two means and significance was assessed using standard statistical methods (ANOVA and *t*-tests), and false discovery rate (FDR) correction for multiple comparisons (Benjamini and Hochberg 1995). All statistics were two-tailed and, where appropriate, included participant sex, age, and IQ as covariates. Effect sizes of between-group differences were calculated using Cohen’s *d*.

### Correlation and mediation analyses

In post-hoc analyses, the interrelationship between regions with impaired activation in Sz (relative to controls) during real- and implied-motion processing was assessed by Pearson’s partial correlation analysis with correction for age, sex, IQ and group membership and correction for multiple comparisons.

Exploratory mediation analyses were conducted within SPSS 26.0 (https://www.ibm.com/products/spss-statistics) using the PROCESS macro (version 4.2) (Hayes 2013). A three-variable path model (model 4) was applied to examine the potential mediating role of MT+ during real- and implied-motion, and bottom-up versus top-down activation patterns. In all models group membership was included as a covariate. The statistical significance of the indirect pathways, reflecting the impact of mediation, was evaluated using a non-parametric bootstrap approach with 10,000 replication samples to obtain a 95% confidence interval (CI) (Preacher and Hayes 2008). The mediation effects were considered statistically significant if the bootstrapped 95% CI did not include zero.

## Results

### Behavioral Measures

As expected, both motion-sensitivity (F(1,43)=9.48, p=.004, *d=1.00*) and ER-40 (F(1,43)=28.39, p<.001, *d=1.63*) scores were significantly lower in Sz relative to NT individuals (**Table 1**).

### Groupwise fMRI

Groupwise activations (p<.01, corrected) elicited by 1) real-motion clips, 2) images with implied-motion and 3) static images with no-motion are shown in **Supplementary Figure 1** superimposed on outlines of the HCP atlas parcels.

When contrasted to no-motion, real-motion stimuli produced significant (p<.05, corrected) activation across all participants within motion-sensitive regions of middle temporal cortex, as well as in lateral-occipital, anterior cingulate, temporo-parietal, superior temporal, and early visual regions. These activations encompassed 9 cortical parcels of the right hemisphere (RH), 4 parcels of the left hemisphere (LH) (**Figure 1A, 1B**) and, subcortically, the PulN of both hemispheres (**Figure 1C, Supplementary Table 1**).

**Figure 1:**
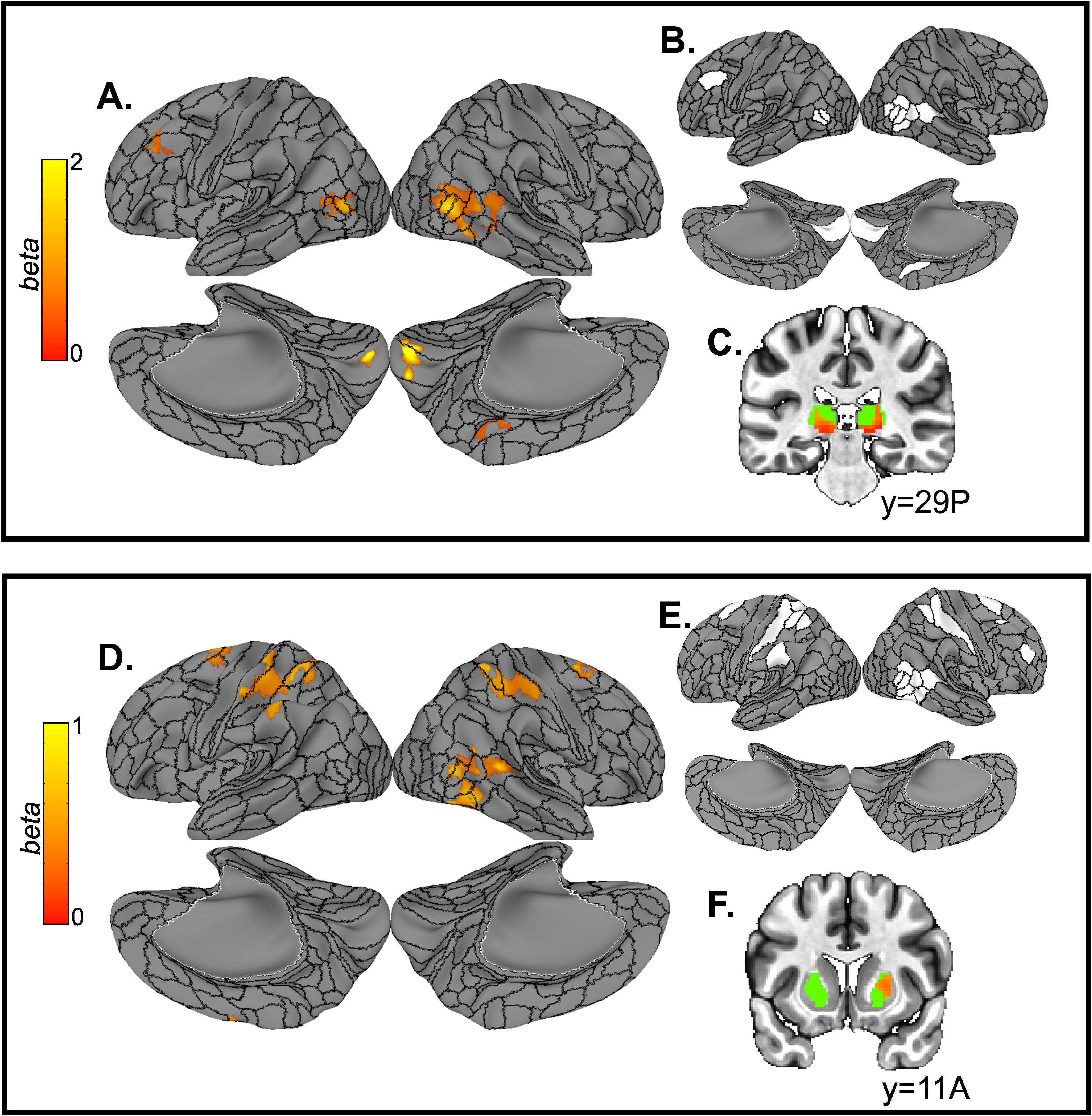
**A)** Cortical regions with significant groupwise activation in the contrast of real-motion versus no-motion. **B)** Corresponding HCP parcels with significant groupwise activation in the real-motion contrast. **C)** Subcortical (pulvinar) groupwise activation superimposed on the anatomical mask used for analysis (green). **D)** Cortical activation elicited by the contrast of implied-motion versus no-motion stimuli. **E)** Corresponding HCP parcels with significant groupwise activation in the implied-motion contrast. **F)** Subcortical (putamen) groupwise activation in implied-motion versus no-motion contrast superimposed on the anatomical mask (green).

The contrast of stimuli with implied-motion versus no-motion stimuli also activated middle temporal regions of the RH across participants. Additionally, a network of fronto-parieto-temporal regions that included portions of the inferior and superior frontal cortex, sensorimotor cortex (SMC), and the posterior STS were also significantly activated. In total, these activations comprised 14 parcels of the RH and 6 of the LH (**Figure 1D, 1E**) as well as the RH putamen nucleus of the basal ganglia (**Figure 1F, Supplementary Table 2**).

### Group Differences

After covarying for participant age, sex, and IQ, both the real-motion (F(1,43)=11.39, p=.002) and implied-motion (F(1,43)=5.65, p=.022) contrasts resulted in significantly reduced activation across all parcels in Sz compared to control participants. In both cases, the group X parcel interaction was also significant (real-motion: F(12,516)=1.84, p=.039; implied-motion: F(19,817)=2.18, p=.003), thus, for each condition post-hoc tests of individual parcels were conducted.

Real-motion contrast:

Compared to NT participants, activation across parcels comprising bilateral area MT+ (MT/MST) (F(1,43)=7.22, p=.010*, d=.85*) was significantly reduced in Sz. The effect of hemisphere (F(1,43)=3.77, p=.060) and the group x hemisphere interaction (F(1,43)=.21, p=.652) were non-significant, therefore, in subsequent analyses, beta parameters were averaged across hemispheres to provide a single MT+ representation.

Activation of parcels comprising pSTS (TPOJ1/TPOJ2) of the RH (F(1,43)=9.29, p=.004, *d=.96*) was also significantly lower in Sz as was activation of bilateral primary visual cortex (V1) (F(1,43)=9.92, p=.003, *d=.84*) (**Figure 2A, 2C, Table 2 top**). For V1, neither the main effect of hemisphere (F(1,43)=.04, p=.834) nor the group x hemisphere interaction (F(1,43)=2.43, p=.126) were significant, thus, mean V1 activation was subsequently used.

**Figure 2:**
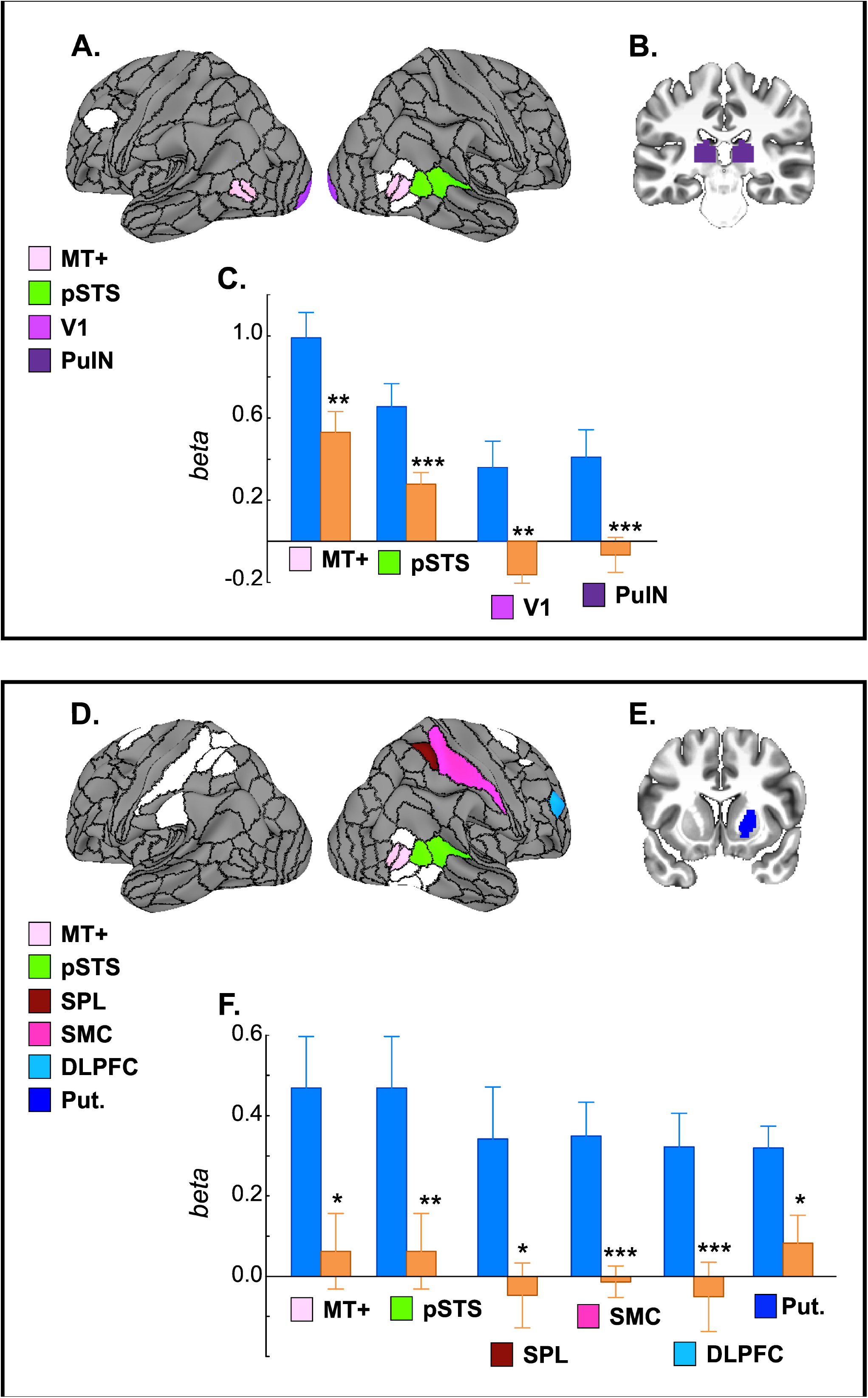
**A**) Cortical parcels (color-coded) with group differences in the real-motion contrast. Parcels in white showed equivalent activation in the Sz and NT groups. **B**) The bilateral pulvinar (PulN) mask used for analyses is in purple. **C**) Bar graphs of mean activation (beta parameter estimates) in the real-motion contrast for NT (blue) and Sz (orange) groups. **D**) Color-coded cortical parcels and **E)** putamen (Put., dark blue) that showed significant group differences in the contrast of implied-versus no-motion **F)** Bar graphs of mean activation in NT and Sz groups within each region. (*, p≤.05; **, p≤.01; ***, p≤.001).

**Table 2:** Mean activation (beta parameter estimate) for NT and SZ elicited by groups within regions with significant group differences during real-motion (top) and implied-motion (bottom) processing. Mean values and statistical significance are given separately for activation elicited by real/implied- and no-motion stimuli as well as for the contrast of real/implied-versus no-motion.

| **Region** | **Real Motion**<br>**(NT/SZ)** | **t(46)** | **p** | **No Motion**<br>**(NT/SZ)** | **t(46)** | **p** | **Real vs. No Motion**<br>**(NT/SZ)** | **t(46)** | **p** |
| --- | --- | --- | --- | --- | --- | --- | --- | --- | --- |
| MT+ | 1.32/1.01 | 2.61 | 0.012 | .33/.49 | -1.41 | 0.166 | .99/.53 | 2.91 | 0.006 |
| V1 | .76/.46 | 2.08 | 0.026 | .40/.62 | -1.32 | 0.263 | .36/-.16 | 2.86 | 0.006 |
| pSTS | .64/.43 | 2.67 | 0.010 | 0.07/0.15 | -1.11 | 0.274 | .65/.28 | 3.26 | 0.002 |
| PulN | .47/.026 | 3.05 | 0.004 | .06/.09 | -0.30 | 0.766 | .41/-.066 | 3.16 | 0.003 |
| **Region** | **Implied Motion**<br>**(NT/SZ)** | **t(46)** | **p** | **No Motion**<br>**(NT/SZ)** | **t(46)** | **p** | **Implied vs. No Motion**<br>**(NT/SZ)** | **t(46)** | **p** |
| MT+ | .80/.55 | 2.03 | 0.048 | .33/.49 | -1.41 | 0.166 | .47/.06 | 2.61 | 0.006 |
| pSTS | .33/.11 | 2.23 | 0.030 | .07/0.15 | -1.11 | 0.274 | .34/-.05 | 3.30 | 0.002 |
| SPL | .20/.01 | 2.13 | 0.038 | -0.14/0.06 | -1.54 | 0.130 | .34/-.05 | 2.69 | 0.010 |
| SMC | .26/.11 | 2.16 | 0.035 | -.09/.12 | -2.62 | 0.011 | .35/-.01 | 4.28 | <.001 |
| DLPFC | .49/.22 | 2.30 | 0.026 | .12/.22 | -0.96 | 0.353 | .32/-.05 | 2.99 | 0.004 |
| Putamen | .22/.07 | 1.80 | 0.070 | -.10/-.02 | -0.96 | 0.343 | .32/.08 | 2.51 | 0.015 |

There were no group differences in the remaining cortical parcels (all p>.4) (**Supplementary Table 1**).

Subcortically, activation in bilateral PulN was significantly lower, overall. in Sz, compared to NT participants (F(1,43)=9.65, p=.003, *d=.93*) with no effect of hemisphere (F(1,43)=.01, p=.916) or group x hemisphere interaction (F(1,43)=.13, p=.717) **(Figure 2B, 2C)**. Mean PulN activation was used in subsequent analyses.

Implied-motion contrast:

Across participants, implied-motion stimuli elicited activation within MT+ parcels of the RH. This activation was reduced in Sz (F(1,43)=5.90, p=.019, *d=.76*) and significantly correlated with MT+ activation deficits during real-motion processing (r_p_=.44 p=.029).

Activation of the pSTS region of the RH was also significantly diminished in Sz (F(1,43)=12.30, p=.001, *d=.97*) and, like MT+, this reduction correlated significantly with the observed pSTS impairments in Sz to real-motion stimuli (r_p_=.62 p=.009).

Additionally, activation of RH parcels comprising dorsolateral prefrontal cortex (DLPFC) (p9-46v; F(1,43)=6.22, p=.017, *d=.82*), primary sensorimotor cortex (SMC) (2; F(1,43)=18.42, p<.001, *d=1.25*) and superior parietal lobe (SPL) (7PC; F(1,43)=6.38, p=.015, *d=.79*) was significantly lower in Sz, compared to NT participants (**Figure 2D, 2F, Table 2 bottom**).

There were no further cortical group differences (all p>.1) (**Supplementary Table 2**). Subcortically, Sz participants showed reduced activation in the putamen of the RH (F(1,43)=4.79, p=.034, *d=.74*) (**Figure 2E, 2F, Table 2 bottom**).

### MT+ intercorrelations

After correcting for multiple comparisons and controlling for participant age, sex, IQ, and group membership, pairwise correlations between regions with group differences during perception of real- and implied-motion were calculated.

For real-motion (**Figure 3A**), activation of MT+ correlated with pSTS (r_p_=.45 p=.002) (**Figure 3B**), V1(r_p_=.38 p=.008) (**Figure 3C**), and PulN (r_p_=.43 p=.004) (**Figure 3D**) activity.

**Figure 3:**
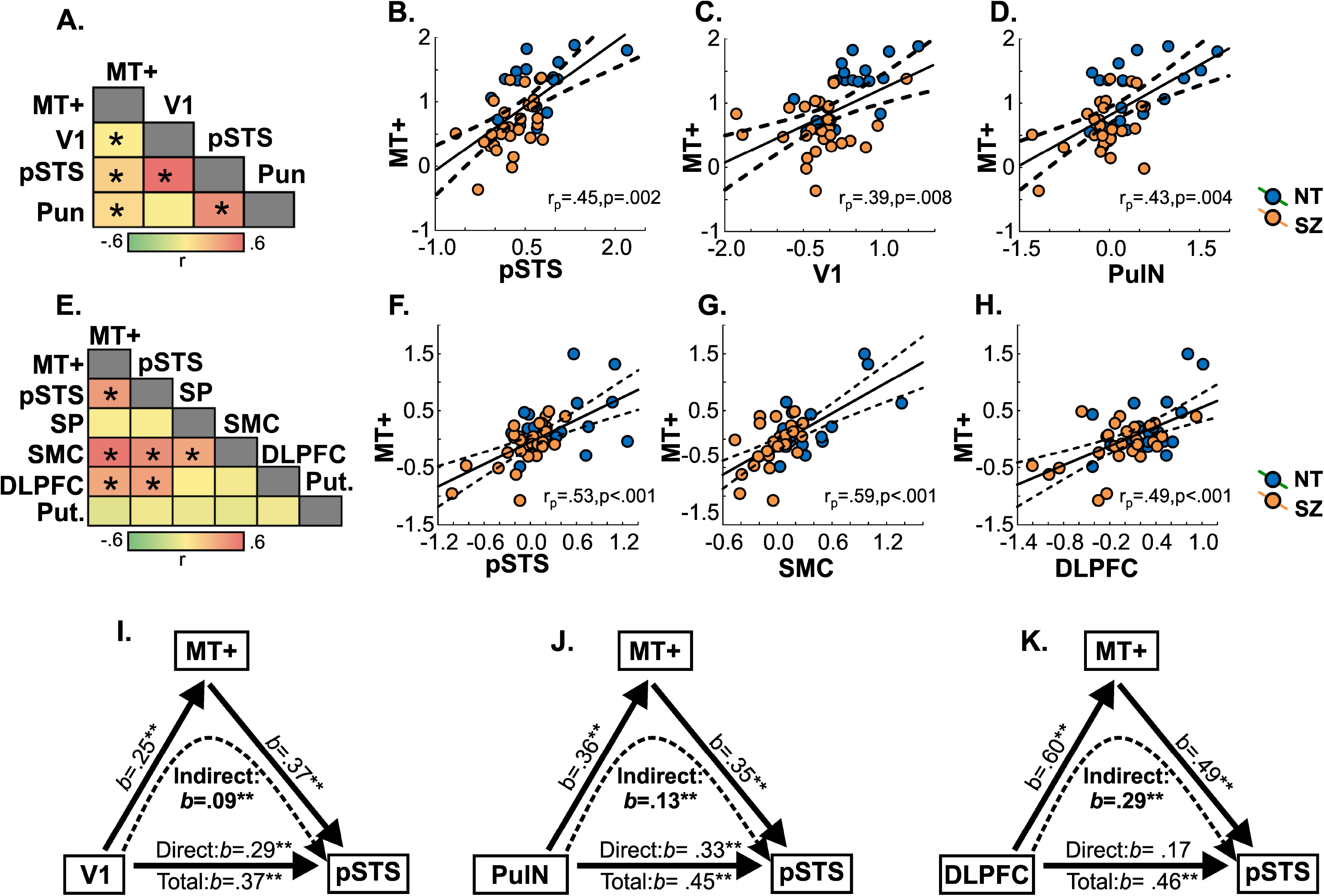
**A)** Cross-correlation between activation in all parcels with group differences in the real-motion contrast. Matrix is of partial correlation coefficients (controlling for group membership, participant age, sex, and IQ). Asterisks denote significant correlation. Scatterplots of mean MT+ activation and activation of **B**) pSTS **C**) V1 and **D**) pulvinar (PulN) for NT (blue) and Sz (orange) participants. **E**) Cross-correlation matrix, as above, for regions with group differences in the implied-motion contrast. MT+ activation correlated with activation of **F**) pSTS, **G**) SMC and **H**) DLPFC. **I**) Schematic representation of mediation models

In the implied-motion contrast (**Figure 3E**), MT+ and pSTS activation were also significantly correlated (r_p_=.53 p<.001) (**Figure 3F**). In addition, MT+ activity was correlated with activation of SMC (r_p_=.59 p<.001) (**Figure 3G**) and DLPFC (r_p_=.49 p<.001) (**Figure 3H**).

A more detailed analysis of these correlations was carried out with exploratory linear mediation analyses designed to evaluate a priori hypotheses regarding pSTS activity, the mediating role of MT+ during real-and implied-motion, and bottom-up versus top-down activation patterns.

Based on the correlation results, four predictor-outcome paths were evaluated, PulN-pSTS and V1-pSTS during real-motion and SMC-pSTS and DLPFC-pSTS during implied-motion. In all models MT+ was used as a potential mediator and sex, age, IQ, and group membership as covariates.

In the real-motion condition, the (indirect) path between both PulN and pSTS (*b*=.13, standard error (SE)=.06, 95% CI=[.03 to .27]) (**Figure 3I**) and V1 and pSTS (*b*=.09, SE=.05, 95% CI=[.01 .20]) (**Figure 3J**) was partially mediated by MT+ (**Supplementary Table 3)**. In both models, all paths were significant, including the direct PulN-pSTS (*b*=.33, SE=.11, 95% CI=[.10 .55]), and V1-pSTS (*b*=.28, SE=.09, 95% CI=[.01 .20]) paths after controlling for MT+ activity, indicating that MT+ acted only as a partial mediator.

By contrast, in the implied-motion condition, the coefficient for the indirect path between DLPFC and pSTS was significant (*b*=.29, SE=.12, 95% CI=[.08 .55], whereas the coefficient for the direct relationship was no longer significant after controlling for MT+ activity (*b*=.17, SE=.13, 95% CI=[-.10 .43]) (**Figure 3K**), consistent with full mediation.

MT+ did not mediate the association between SMC and pSTS activity (indirect path: *b*=.22, SE=.21, 95% CI=[-.24 .59]).

### Behavioral correlations

Across participants, there was a significant partial correlation between behavioral measures of motion-sensitivity and MT+ activation to real-motion stimuli (r_p_= .39, p=.008). This relationship was independently significant in the Sz group (r_p_= .46, p=.020) such that reduced MT+ activity predicted poor motion sensitivity (**Figure 4A**).

**Figure 4:**
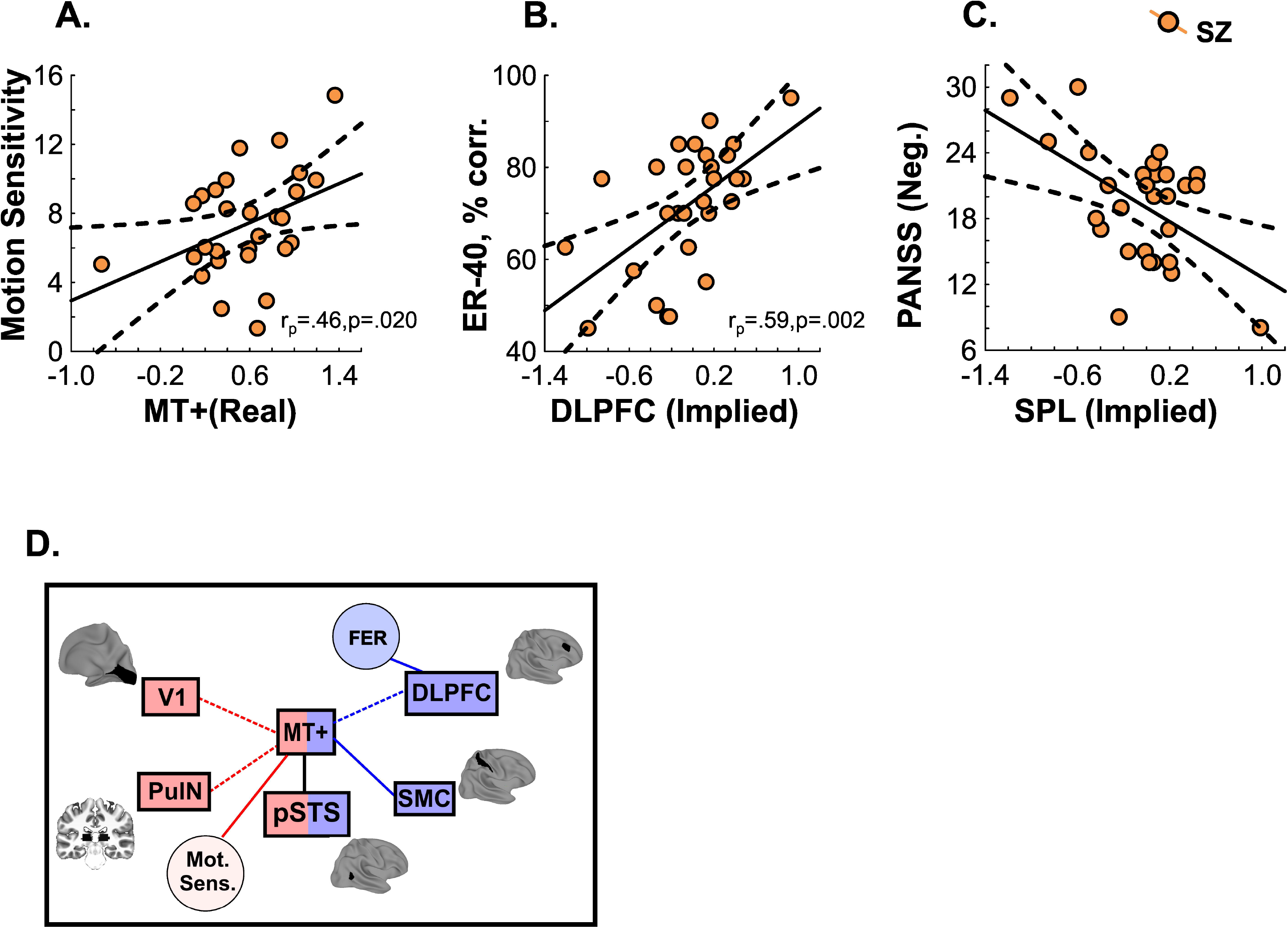
**A**) Correlation between MT+ activation in Sz during real-motion processing and motion-sensitivity score (1/threshold) derived from coherent motion detection paradigm. **B)** Correlation of DLPFC activation to implied-motion stimuli and performance (percent correct) on ER-40 test of face emotion recognition. **C)** Schematic representation of the interrelationship between behavioral measures of face emotion recognition (FER) and motion sensitivity (Mot. Sens.) and cortical/subcortical activation associated with real-(red) and implied-motion (blue) perception. Dashed lines indicate putative paths from mediation analyses between V1/PulN and MT+ during real-motion and DLPFC and MT+ during implied-motion. MT+ and pSTS were commonly activated by real and implied-motion stimuli and, in both cases, were strongly linked with one another suggesting a possible bridging role for MT+.

Additionally, across participants (r_p_= .54, p<.001), and in Sz alone (r_p_= .59, p=.002), poor FER (reduced ER-40 scores) were predicted by reduced DLPFC activation during implied-motion perception (**Figure 4B**).

Lastly, in Sz, symptom severity as measured by the PANSS negative syndrome scale correlated with impaired SPL activation (r_p_= -.52, p=.006) (**Figure 4C**).

A schematic summary of the interrelationship between behavioral and fMRI measures of real- and implied-motion perception is shown in **Figure 4D**.

## Discussion

Impaired motion perception is one of the most replicated examples of visual dysfunction in Sz (Chen 2011; Hong et al. 2005; Kim et al. 2006; Li 2002; Martinez et al. 2018) and has been shown to involve reduced activation of visual sensory regions, including MT+ (Chen et al. 2008; Lencer et al. 2005; Martinez et al. 2018).

In recent years, there has been increasing appreciation of the link between the visual motion system and social brain networks (Guterstam and Graziano 2020; Pavlova 2012). In particular, it has been hypothesized that MT+ forms a bridge between visual-sensory and high-order cognitive regions (Yeo et al. 2011). Here, we tested this hypothesis in Sz in regard to the MNS, which supports mentalization operations that assist in interpreting visual input, and which has been implicated in neurocognitive dysfunction across neuropsychiatric disorders. We used naturalistic stimuli manipulated to create real-motion, implied-motion, and no-motion percepts to directly compare systems engaged by “bottom-up” versus “top-down” sources of motion information.

Compared to controls, individuals with Sz showed reduced MT+ activation to both real- and implied-motion stimuli, as well as reduced pSTS activation (Duarte et al. 2022). While MT+ is not considered a core MNS region, several studies have documented its involvement within the extended MNS system (Caspers et al. 2010; Molenberghs et al. 2012). Here, we provide the first demonstration that, in Sz, MT+/pSTS activation is impaired during implicit-as well as explicit-motion perception, with a strong correlation between the two sets of deficits. These findings support concepts of MT+ as a bridge between purely sensory and higher-order cognitive dysfunction, and thus as a potential specific target for therapeutic intervention.

*Real-motion:* In addition to MT+ and pSTS, the contrast of real-versus no-motion resulted in robust activation of subcortical (PulN) and early visual cortex (V1) both of which were tightly intercorrelated with impaired MT+ activation. Mediation analyses suggested that the association between PulN-pSTS and V1-pSTS was partially mediated by MT+ activity, suggesting that impaired real-motion processing in Sz may be, at least in part, due to dysfunctional bottom-up input to MT+. We have previously documented reduced PulN and V1 activity in Sz in response to simple motion stimuli (Martinez et al. 2019) and dynamic emotional faces (Martínez et al. 2022), we now show that these deficits extend as well to naturalistic moving scenes. Finally, in Sz, we observed intercorrelated reductions in motion-perception sensitivity, as measured behaviorally, and activation of MT+, consistent with our prior investigation (Martinez et al. 2018).

*Implied-motion*: As in previous investigations (Fawcett et al. 2007; Kourtzi and Kanwisher 2000; Krekelberg et al. 2005; Proverbio et al. 2009; Senior et al. 2000) MT+ showed greater activation when participants viewed static images with implied-motion cues than when viewing static images that implied the absence of motion. As in the case of real-motion, MT+ activation during implied-motion perception was also reduced in Sz. Unlike real-motion, however, which was associated with processes engaging subcortical and early visual regions, MT+ activation to implied-motion stimuli correlated significantly with activation of sensorimotor and prefrontal cortex. These findings support the hypothesis that the involvement of MT+ in implied-motion perception is the result of its interaction with higher-level cortical regions (Kourtzi and Kanwisher 2000; Lorteije et al. 2006; Senior et al. 2000), and suggests that, in Sz, these (top-down) modulatory interactions are abnormal.

Several cortical and subcortical nodes of the human MNS (Mineo et al. 2018; Proverbio et al. 2009; Urgesi et al. 2006) were also activated during perception of implied-motion, though Sz individuals showed reduced activity in only a subset of these including the superior parietal lobe, primary sensorimotor and DLPFC, as well as putamen. Activation of other MNS regions, most notably premotor and supplementary motor nodes, were not significantly different between groups, suggesting that the observed deficits do not reflect global malfunction of the MNS, but rather impairment of subcomponents that may contribute differentially to visual perceptual functioning in Sz.

In the present study, impaired prefrontal (DLPFC) activation during implied-motion bridged between reduced MT+ activity and FER accuracy in Sz. These findings parallel our prior observations that when a simple visual motion stimulus was used to elicit MT+ activation, MT+ activity correlated directly with FER accuracy (Martinez et al. 2018), but when a dynamic emotional face stimulus was used, pSTS served as the intervening region (Martínez et al. 2022). The MNS and superior temporal regions are known to play synergistic roles in FER (Bonini et al. 2022). Our findings demonstrate that both may interact with MT+ to process the explicit and/or implied-motion information needed for accurate FER determination, suggesting that developmental disruptions in MT+ organization may undermine processing across cortical networks. If confirmed, our findings argue for greater focus on MT+ contributions to social cognitive processing, in addition to its well-documented role in visual motion processing.

Lastly, dysfunctional superior parietal activation was associated with greater negative symptoms in the patient group, in agreement with prior studies (Thakkar et al. 2014) and in support of the hypotheses that a dysfunctional MNS may underlie some cognitive deficits in Sz.

*Third visual pathway*: For over 30 years, the prevailing model of visual system functional organization has proposed the existence of two segregated visual streams (Goodale and Milner 1992), the dorsal (“where”) and ventral (“what”) pathways. Recently, it has been proposed that a third visual pathway leading from V1 to MT+ and pSTS is a functionally distinct pathway that differentially processes information relevant to social cognition (Pitcher and Ungerleider 2021). The present study adds to an accumulating literature demonstrating dysfunction of this system in Sz independent of the previously described dorsal/ventral streams. For example, we have previously observed deficits in V1 and MT+ activation to simple motion stimuli (Martinez et al. 2019), and in V1, MT+ and pSTS to moving faces (Martínez et al. 2022) in the absence of impairments within ventral and dorsal stream regions. Our present findings of impaired V1, MT+ and pSTS to real-motion, despite limited engagement of dorsal visual cortex, provide further evidence of dysfunction within the proposed third visual pathway, independent of dysfunction within the canonical visual streams.

Further, while dorsal stream deficits in Sz are largely observed to stimuli that preferentially engage the subcortical magnocellular pathway and reflect primarily bottom-up, subcortically-driven, dysfunction (Javitt and Freedman 2015), our present findings suggest that third visual pathway (MT+/pSTS) dysfunction may be observed even under circumstances of only top-down input. One potential explanation for the finding is that both bottom-up and top-down inputs are independently impaired (Scheliga et al. 2022). Alternately, the failure of bottom-up input over time may lead to a loss of precision within convergent motion processing modules in MT+ that respond to both real- and implied-motion (Krekelberg et al. 2005), which then undermines their ability to participate within higher cortical networks.

Finally, we have previously observed that motion-processing deficits predate illness onset in Sz and may predict conversion to psychosis (Martinez et al. 2018). Future studies within the clinical high-risk population are needed to evaluate the ontogeny of bottom-up versus top-down motion processing impairments. Furthermore, given the increasing focus on social cognition deficits as a critical determinant of functional outcome not only in Sz (Green et al. 2019; Javitt 2023) but also in disorders such as autism (Sasson et al. 2020), future studies on the causes and consequences of third visual pathway dysfunction are warranted.

Several limitations of this study must be acknowledged. First, we showed correlations between prefrontal activation and ER-40, a normed task for the assessment of FER. In contrast, no normed tasks were available for the detection of implied-motion. Future studies with a wider array of stimuli and comprehensive behavioral measures are warranted. Second, while deficits did not correlate with medication dose, all Sz participants were receiving antipsychotics which may have affected the between-group results. Finally, although differences between groups were statistically robust, they require replication in larger samples.

## Conclusions

Motion-processing deficits are a hallmark of Sz linked to impaired processing within the cortical and subcortical magnocellular/dorsal visual pathway and with impairments in higher-order social cognitive processes. To date, deficits in MT+ activation have been investigated primarily with simple stimuli such as moving dots displays. Here, we show that MT+ deficits are also observed when motion is embedded within more complex scenes and depends upon impaired bottom-up visual input through V1 and PulN. Additionally, during implied-motion perception MT+ deficits in Sz were associated with a wider network involving sensorimotor and prefrontal nodes of the MNS. The correlated deficits in MT+ activation to real- and implied-motion processing argue for a critical role for MT+ in bridging between bottom-up and top-down processes during visual perception in Sz.

## Data Availability

All data produced in the present study are available upon reasonable request to the authors.

## Acknowledgements

The authors thank the Clinical Research and Evaluation Facility at NKI and all research participants for their contributions. This work was supported by National Institute of Mental Health (grant number MH49334).

## Notes

### Competing Interest Statement

The authors have declared no competing interest.

### Funding Statement

This study was funded by by National Institute of Mental Health (grant number MH49334).

### Author Declarations

Ethics committee/IRB of the Nathan Kline Institute for Psychiatric Research gave ethical approval for this work.

