## Supplementary Material for "Bottom-up and top-down contributions to impaired motion processing in schizophrenia"

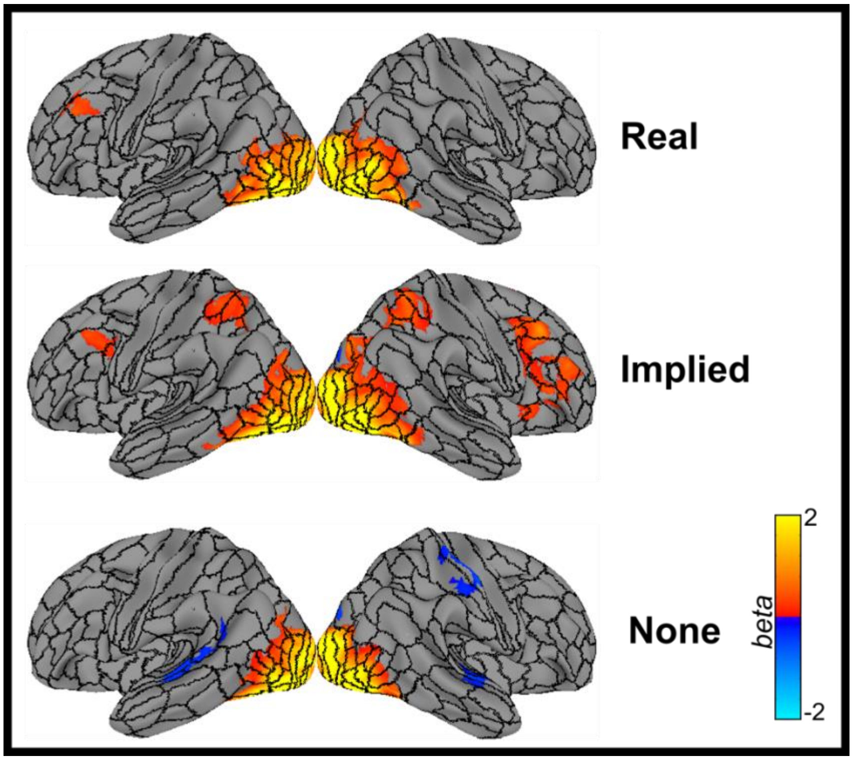


*<u>Supplementary Figure 1:</u>* Mean activation (across all participants) elicited by real-motion (**top**), implied-motion (**middle**) and no-motion stimuli (**bottom**) superimposed on outlines of the HCP atlas parcels.

| Parcel | Mean beta *(NT/SZ)* | *t(46)* | *p* | Cohen's *d* |
|---|---|---|---|---|
| V1 (LH) | .29/-.16 | 2.57 | *0.036* | 0.76 |
| V1 (RH) | .54/-.06 | 3.61 | *0.034* | 1.06 |
| LO3 (RH) | .46/.19 | 1.36 | 0.250 | 0.40 |
| MT (LH) | .95/.44 | 2.27 | *0.028* | 0.66 |
| MT (RH) | 1.10/.52 | 3.80 | *0.006* | 1.11 |
| MST (LH) | .95/.45 | 1.97 | 0.054 | 0.58 |
| MST (RH) | 1.3/.72 | 3.73 | *0.005* | 1.09 |
| FST (RH) | .73/.60 | 0.91 | 0.440 | 0.27 |
| TPOJ1 (RH) | .71/.26 | 2.61 | *0.034* | 0.77 |
| TPOJ2 (RH) | .65/.30 | 2.77 | *0.034* | 0.81 |
| TPOJ3 (RH) | .47/.40 | 0.51 | 0.613 | 0.15 |
| 5mv (RH) | .35/.19 | 0.98 | 0.447 | 0.29 |
| 8c (LH) | .11/.06 | 0.37 | 0.712 | 0.11 |
| PulN (LH) | .42/-.02 | 2.02 | 0.111 | 0.59 |
| PulN (RH) | .41/-.11 | 3.49 | *0.009* | 1.02 |

*Supplementary Table 1*: All parcels with significant activation across all participants during real motion-perception. Activation of the left (LH) and right (RH) hemisphere pulvinar (PulN) is given at bottom. Mean activation (beta parameter) for the control (NT) and schizophrenia (SZ) groups is provided. Statistical (t and p) values and effect sizes (Cohen's d) are for the NT versus SZ comparison.

| **Parcel** | **Mean beta *(NT/SZ)*** | ***t(46)*** | ***p*** | **Cohen's *d*** |
|---|---|---|---|---|
| MT (RH) | .19/-.17 | 2.62 | *0.041* | *0.78* |
| MST (RH) | .30/-.08 | 3.10 | *0.035* | *0.91* |
| FST (RH) | .39/.10 | 0.91 | 0.142 | 0.56 |
| PH (RH) | .27/.35 | 0.27 | 0.711 | -0.15 |
| PHT (RH) | .25/.23 | 0.02 | 0.895 | 0.04 |
| TPOJ1 (RH) | .26/-.08 | 2.42 | *0.048* | *0.71* |
| TPOJ2 (RH) | .34/-.03 | 3.00 | *0.030* | *0.88* |
| TPOJ3 (RH) | .26/.19 | 0.56 | 0.641 | 0.22 |
| PF (LH) | .20/.13 | 0.38 | 0.671 | 0.18 |
| IP0 (RH) | .25/.20 | 0.16 | 0.768 | 0.12 |
| LIPv (LH) | .28/.07 | 1.52 | 0.286 | 0.44 |
| 7AL (LH) | .19/.04 | 1.20 | 0.450 | 0.35 |
| 7PC (LH) | .29/.18 | 0.66 | 0.672 | 0.19 |
| 7PC (RH) | .34/-.05 | 2.69 | *0.042* | *0.79* |
| 2 (LH) | .22/.10 | 1.20 | 0.415 | 0.35 |
| 2 (RH) | .35/-.01 | 4.28 | *0.002* | *1.25* |
| 6ma (LH) | .29/.13 | 0.96 | 0.552 | 0.28 |
| 6ma (RH) | .10/.17 | -0.80 | 0.638 | -0.24 |
| i6-8 (RH) | .23/.21 | 0.18 | 0.900 | 0.05 |
| p9-46v (RH) | .33/-.01 | 3.08 | *0.038* | *0.82* |
| Put. (RH) | .32/.08 | 2.52 | 0.048 | *0.74* |

*Supplementary Table 2*: Same as above for the implied motion condition. Subcortically, the putamen (Put.) of the RH was significantly activated.

A.

| | | Path | b | SE | t | p | 95% CI LL | 95% CI UL |
|---|---|---|---|---|---|---|---|---|
| | PulN-MT+ | a | 0.36 | 0.11 | 3.34 | 0.002 | 0.14 | 0.58 |
| *X=PulN* | MT+-pSTS | b | 0.35 | 0.14 | 2.54 | 0.015 | 0.07 | 0.63 |
| *Y=pSTS* | PulN-pSTS | c | 0.45 | 0.11 | 4.28 | <.001 | 0.24 | 0.67 |
| *M=MT+* | PulN-pSTS I MT+ | c' | 0.33 | 0.15 | 2.93 | 0.005 | 0.10 | 0.55 |
| | Indirect | a*b | 0.13 | 0.06 | -- | -- | 0.01 | 0.55 |

B.

| | | Path | b | SE | t | p | 95% CI LL | 95% CI UL |
|---|---|---|---|---|---|---|---|---|
| | V1-MT+ | a | 0.25 | 0.09 | 2.62 | 0.012 | 0.06 | 0.43 |
| *X=V1* | MT+-pSTS | b | 0.37 | 0.13 | 2.88 | 0.006 | 0.11 | 0.64 |
| *Y=pSTS* | V1-pSTS | c | 0.38 | 0.09 | 4.30 | <.001 | 0.20 | 0.55 |
| *M=MT+* | V1-pSTS I MT+ | c' | 0.29 | 0.09 | 3.27 | 0.002 | 0.11 | 0.46 |
| | Indirect | a*b | 0.09 | 0.05 | -- | -- | 0.01 | 0.20 |

C.

| | | Path | b | SE | t | p | 95% CI LL | 95% CI UL |
|---|---|---|---|---|---|---|---|---|
| | DLPFC-MT+ | a | 0.60 | 0.11 | 5.25 | <.001 | 0.37 | 0.83 |
| *X=DLPFC* | MT+-pSTS | b | 0.49 | 0.13 | 3.63 | <.001 | 0.22 | 0.76 |
| *Y=pSTS* | DLPFC-pSTS | c | 0.46 | 0.12 | 3.99 | <.001 | 0.23 | 0.69 |
| *M=MT+* | DLPFC-pSTS I MT+ | c' | 0.17 | 0.13 | 1.30 | 0.199 | -0.09 | 0.43 |
| | Indirect | a*b | 0.29 | 0.12 | -- | -- | 0.08 | 0.55 |

*Supplementary Table 3*: Results of mediation analyses testing whether a proposed causal effect of X (predictor) on Y (outcome) may be transmitted through a mediating (M) variable. For each mediation analysis, the coefficients (b), standard error (SE), t-statistic, p-value and lower (LL) and upper (UL) levels for the 95% confidence interval (CI) are given. Standardized beta coefficients are given for the direct paths: (a) X and M; (b) M and Y; (c) X and Y; (c') X and Y, conditional on M and for the indirect (a*b) effect. All models controlled for group membership. **A**) In the first model, the association between PulN and pSTS was partially mediated by MT+ activation during real-motion. **B**) Similarly, MT+ activation partially mediated the association between V1 and pSTS in the real motion condition. C) During implied-motion processing, the relationship between DLPFC and pSTS was fully mediated by MT+.
